# Interpreting Changes in Life Expectancy During Temporary Mortality Shocks

**DOI:** 10.1101/2022.03.17.22272583

**Authors:** Patrick Heuveline

## Abstract

Life expectancy is the most popular mortality indicator with demographers. Unless specified otherwise, it implicitly refers to the value *at birth* (age 0) of one of the functions derived through a *period* life table, a key tool of demographic and actuarial analysis. Demographers tend to favor life expectancy because it is a *pure* measure of the mortality conditions faced by a population, unaffected by that population’s age structure. Life expectancy also has an intuitive interpretation, conditional on the assumption that mortality conditions remain unchanged, as the expected age at death of an average newborn. If life table construction might be limited to an inner circle of demographers and actuaries, this interpretative ease gives life expectancy a much broader appeal.

---

Demographers also use life expectancy to track mortality trends. The United Nations Population Division, for instance, estimates that global life expectancy declined by 1.74 years between 2019 and 2021 (United Nations 2022). This decline provides a measure of the direct and indirect impact of COVID-19 pandemic on survival chances. This measure still has the advantage of being unaffected by the population’s age structure. As long time series of life expectancy values are available, estimating changes in life expectancy also allows for comparisons of the pace of mortality change over time (Aburto et al. 2021; Heuveline 2022). The interpretation of particular values of changes in life expectancy is no longer straightforward, however, because our usual, intuitive interpretation of life expectancy assumes no change in mortality. Simply put, there would seem to be an inherent contradiction in measuring mortality change with a metric best understood under the assumption that mortality does not change.

What then is this 1.74-year-decline a measure of? In this article, I argue that this specific value can still be interpreted, but as a measure of premature mortality in a death cohort (those who died in 2021) rather than as a measure of the change in longevity between the 2019 and 2021 birth cohorts. To clarify this, I return to the two basic representations of the period life table, as a synthetic cohort and as a stationary population. The common interpretation of life expectancy derives from the former, where life table functions represent the future experience of a *synthetic* cohort, an imaginary cohort experiencing at every age the mortality conditions experienced during a reference period by population members of that age. Under temporary mortality shocks—like the Spanish flu, wars, or Covid-19—the alternative interpretation of the difference in life expectancy proposed here can be derived by taking the life table functions to represent instead the characteristics of a stationary population.

## The period life table as a synthetic cohort

A cohort life table represents the complete survival and eventual extinction of a cohort (typically a birth cohort). In a cohort life table, life expectancy at birth is “the sum of all person-years lived by the cohort divided by the original number in the cohort” (Preston, Heuveline, and Guillot 2001). The number of years lived that each member of the cohort contributes to the sum is their length of live, that is, their age at death. And since in a closed cohort (no migration), the original number in the cohort is the number of deaths, the cohort life expectancy at birth is simply the average age at death of cohort members. In continuous notation, this can be expressed as:

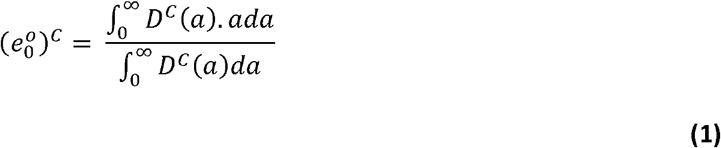

where (*e*_0_°)^c^ is the cohort life expectancy at birth and *D*^c^ (*a*) represents the number of cohort members dying at age *a* (and thus living exactly *a* years) in a closed cohort.

Using the same approach, a period life table describes what would happen to a hypothetical (“synthetic”) cohort if its members were subjected for their entire lives to the mortality conditions of a period, which are typically operationalized as a set of age-specific death rates for that period (Preston, Heuveline, and Guillot 2001). In continuous notation, the period life expectancy at birth can similarly be expressed as:

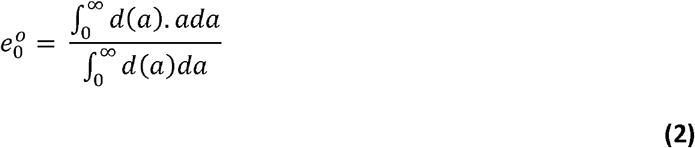

where *e*_0_° is the period life expectancy at birth and *d*(*a*) represents the number of decrements at age *a* in the life table. Starting with an arbitrary number of cohort members at birth (the “radix” of the life table), the construction of the period life table aims to generate the number of survivors to any age *a, l*(*a*), resulting from the lifetime exposure to the age-specific mortality rates *μ*(*x*) from birth to age *a*:

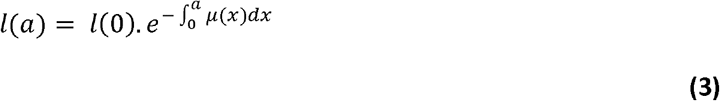

The distribution of deaths, *d*(*a*), is obtained by multiplying this distribution of survivors, *l*(*a*), by the age-specific death rate at the corresponding age, *μ*(*a*):

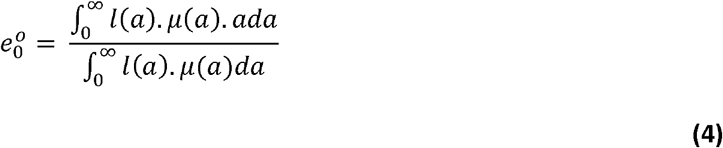

The radix of the life table is but a scaling factor that affects the numerator and the denominator of equation (4) in equal measures. Combining equations (3) and (4), period life expectancy could be expressed purely as a function of the age-specific mortality rates *μ*(*a*) from birth to a maximum age. It represents the average age at death among members of the synthetic cohort subjected throughout their life to these rates, in that sense a “pure” measure of mortality conditions during that period, independent of other population characteristics such as its age distribution. In a probabilistic framework, life expectancy at birth represents the “expected” value of the length of life of a member of the synthetic cohort—a newborn subjected throughout their entire lifetime to the mortality rates *μ*(*a*). The very term life expectancy is closely tied to this “forward-looking” interpretation.

The fact that life expectancy only depends on the age-specific mortality rates is a desirable property in comparisons of mortality conditions across or within populations, or across periods. In some cases, the difference between two life expectancies can still be interpreted in a probabilistic, forward-looking framework. The difference may represent, for instance, how much longer a person could be expected to live under the mortality conditions experienced by one population, or one sub-population, compared to those experienced by another population, or sub-population, were the mortality conditions of these two populations or sub-populations to remain unchanged throughout this person’s lifetime. The difference is then often referred to as the *gap* in life expectancy resulting from one populations or sub-population experiencing higher mortality than another population or sub-population (e.g., ethnic/racial gap in life expectancy).

The basis for Nathan Keyfitz (1977) seminal paper on changes in life expectancy, another example would be the difference between life expectancy before and after a hypothetical medical improvement that would eliminate a cause of death. Within this framework, however, the interpretation of the difference in life expectancy becomes more challenging when what separates the mortality conditions being compared is a temporary phenomenon (mortality “shock”). The difference between pre-pandemic life expectancy and life expectancy during the pandemic, for instance, compares a counterfactual expected age at death had the pandemic never occurred (and had mortality from other causes not changed either) with a hypothetical age at death if pandemic mortality continues throughout a person’s lifetime. Perhaps not entirely impossible with the periodic emergence of new variants, the latter, hypothetical scenario is nonetheless relatively unlikely. Independent from the population age structure, this difference in life expectancy still provides a useful measure of the temporary impact of the pandemic but can no longer be interpreted as a difference in expected ages at death before and during the pandemic.

## The period life table as a stationary population

The second interpretation of the period life table is that its functions provide the characteristics of a specific, hypothetical population: the current population’s “stationary equivalent.” This population’s “stationary equivalent” refers to the stationary population that would emerge if the population indefinitely experienced the mortality conditions of the reference period, with no migration and a constant number of births per unit of time. Among these population characteristics, the number of survivors to age *a, l*(*a*), provides the age distribution of the stationary equivalent population and life expectancy indicates the mean age at death in that stationary equivalent population.

The mean age at death in the actual population at a given time can be expressed as:

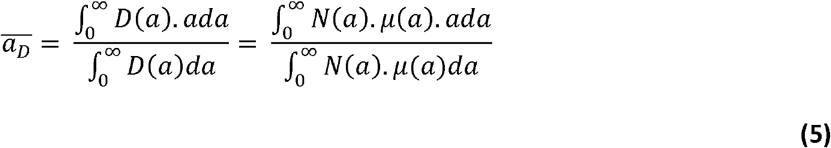

where *D*(*a*) is the number of persons dying at exact age *a* and *N*(*a*) is the number of persons of exact age *a* in the population (with reference to time omitted to lighten notations). Comparing equations (4) and (5), life expectancy at birth appears as an age-standardized form of the mean age at death in the sense that it substitutes the actual population distribution by age, *N*(*a*), with the age distribution in the stationary equivalent population, *l*(*a*).

This standardization differs from the most common form of standardization, which involves the substitution of the actual population distribution with an *external* standard distribution. By comparison, the form of standardization life expectancy entails can be described as an *internal* standardization to the extent that the age distribution being used, *l*(*a*), derives instead from the population’s own age-specific mortality rates. When comparing demographic indicators in two populations, the goal of standardization is to remove the influence of their actual age distributions, which both forms of standardization achieve. The main difference here is that internal standardization does depend on the actual mortality conditions faced by the population (since the distribution *l*(*a*) is derived from the age-specific mortality rates *μ*(*a*)), whereas external standardization does not.

For some analyses, the fact that the comparison of two “internally standardized” populations involves applying potentially different population distributions to the two populations being compared might be perceived as a disadvantage (Modig, Rau, and Ahlbom 2020). However, internal standardization is immune to one issue potentially encountered with external standardization. If population *A* has twice the mortality rates of population *B* at every age, an externally standardized mean age at death would be the same in the two populations. This is counterintuitive as one would expect an average person to die younger in population *A*. The intuition is often correct, because higher mortality would typically (albeit not necessarily) induce a “younger” age distribution in population *A* than in population *B*. While analysts often treat age distribution as a demographic parameter whose influence needs to be purged to obtain a pure measure of the phenomenon of interest, here mortality, age distribution reflects the population’s history of fertility, mortality, and migration. While external standardization removes all traces of this demographic history, the internal standardization life expectancy performs only removes those related to fertility and migration but maintains some coherence between current mortality levels and the age structure used as internal standard. Under these conditions, life expectancy in population *A* would indeed be lower than in population *B*.

Moreover, there are several situations in which an alternative interpretation of life expectancy as a stationary-equivalent indicator can usefully complement its more common probabilistic, forward-looking interpretation. First, this alternative interpretation applies equally well to very short reference periods. When mortality conditions change rapidly, one may want to track these changes with high frequency, for periods shorter than a full year (e.g., Trias-Llimos Riffe, and Bilal 2020; Ghislandi et al. 2022). Due to the seasonality of mortality conditions, however, the probabilistic interpretation of life expectancy for reference periods shorter than a full year implies an implausible lifetime experience, looping conditions in some seasons while skipping conditions in other seasons. Ho and Noymer (2017) refer to such period life expectancies as “pseudo seasonal” expectancies. Second, as described above, the probabilistic interpretation is problematic when considering differences in life expectancy resulting from temporary mortality shocks.

If life expectancy can be interpreted as an internally standardized average age at death, what is the corresponding interpretation of a difference in life expectancy? Re-writing Pollard’s (1988) formula, the difference in life expectancy appears as follows (see Appendix Part 1 for a derivation of this equation and its relation to Keyfitz’ (1977) hypothetical change mentioned above):

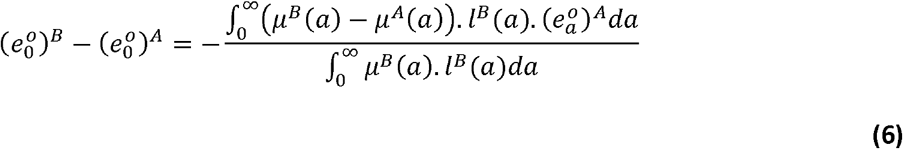

where superscripts *A* and *B* refer to two different life tables (corresponding to different populations or sub-populations, or to the same population in two different periods). Extending the stationary equivalent interpretation of the life table, the absolute value of the (negative) difference in life expectancy shown in equation (6) can be seen as the internally standardized form of the Mean Unfulfilled Lifespan (*MUL*), a measure of premature mortality (Heuveline 2021):

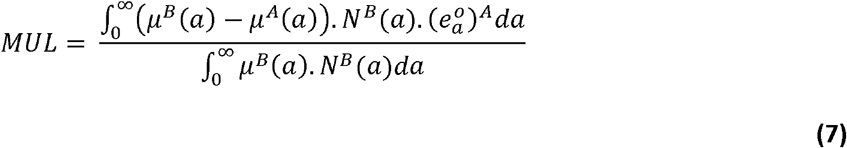

Again, the only difference between the two ratios is that in the difference in life expectancy, as shown in equation (6), the distribution of survivors by age in the period life table *B*, replaces the actual age distribution of population *B, N*^*B*^(*a*), used to calculate the *MUL*, as shown in equation (7).

To interpret the *MUL*, and its stationary equivalent, the difference in life expectancy, first note that when mortality rates at age *a* are higher in period life table *B* than in period life table *A*, the first product in the numerator’s sum represents “excess deaths” at age *a, D*^*E*^(*a*). This number is defined as the difference between the actual number of deaths at age *a* in population or period *B, D*^*B*^(*a*), and a counterfactual number of deaths in population *B* had the (typically lower) mortality rate of population or period *A* prevailed instead at that age:

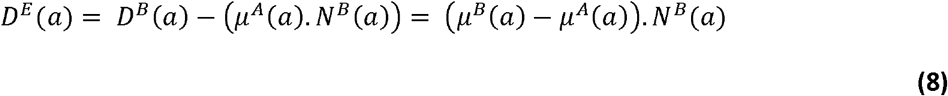

Each excess death at age *a* corresponds to a person who died at that age in population or period *B* and who would have lived in population or period *A*, and the number of additional years this person would have been expected to live is the life expectancy at age *a* in population or period *A*, (*e*_a_^*o*^)^A^. This life expectancy thus represents the number of years of life lost by that person due to the mortality differences between the two populations or periods. Summing across all excess deaths, the total years of life lost (*YLL*) to excess mortality (i.e., to the difference in mortality between the two populations or periods) is:

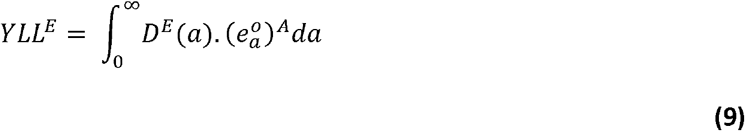

which is the numerator of the *MUL*. Meanwhile, the sum in its denominator adds up to all the deaths (of all ages) in population or period *B, D*^*B*^. Averaging *YLL*^*E*^ over the total number of deaths, the *MUL* thus represents the average number of years of life lost to excess mortality per death (irrespective of cause of death) in population or period *B*:

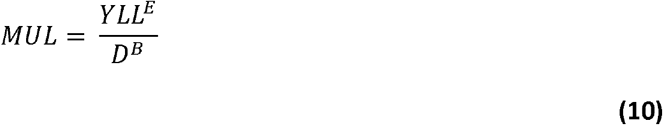

The *MUL* can be related to several other indicators of premature mortality (see Appendix Part 2). One of these relationships involves the *P-score*, the ratio of excess to expected deaths in the absence of mortality changes, and the average number of years of life lost to excess mortality per *excess* death:

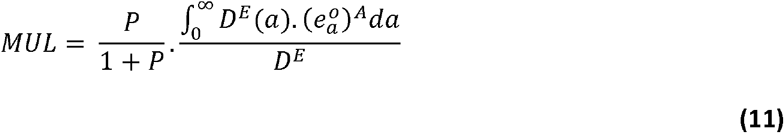

Equation (11) provides intuition for the value of the *MUL*. This value is the product of (1) the proportion of all deaths in population or period *B* that can be considered excess deaths by comparing the prevailing mortality conditions in population or period *B* to the more favorable mortality conditions in population or period *A* (the ratio *P*/1+*P*) and of (2) how much longer on average a person who died due to the difference in these mortality conditions would have been expected to live under the more favorable conditions in population or period *A*.

As an illustration, Karlinsky and Kobak (2022) estimate that 511,232 of the 3,455,604 deaths in the United States in 2021 (14.8%) are excess deaths that would not have occurred under pre-pandemic mortality conditions. Meanwhile, Goldstein and Lee (2020) estimate that those who die of Covid-19 have on average 11.7 years of remaining life expectancy. This number likely increased in 2021 relative to 2020 as the age-structure of Covid-mortality shifted towards younger ages as a result of higher vaccination rates among the elderly. As Covid-19 represents the bulk of excess mortality in the United States in 2021 relative to 2019, this suggests the US *MUL* was at least 1.73 years in 2021.

The value of the *MUL* depends on demographic characteristics other than mortality, such as the population age distribution. All else equal, the average number of years of life lost to excess mortality per excess deaths takes larger values in populations with younger age distributions, because the age distribution of excess deaths itself shifts toward younger ages at which the values of life expectancy are larger. The value of the *P-score* also depends on the population age structure (except in special cases such as when the relative difference in age-specific mortality rates between the prevailing and more favorable conditions is the same at all ages). As life expectancy with respect to the mean age at death, by substituting the stationary equivalent distribution to the actual population distribution the absolute value of the difference in life expectancy then provides a useful internal age-standardization of the value of the *MUL*. To return to the example of the United States, the 2019-21 decline in life expectancy at birth has been estimated at 1.9 years (Heuveline 2022). This is larger than the value of the *MUL* approximated above, consistent with the fact that the current US age distribution is “older” than its stationary-equivalent due to the aging of the large birth cohorts of the baby-boom years.

An indicator of premature mortality, the *MUL* is typically positive when comparing prevailing mortality conditions to more favorable ones in the past, which implies that the difference in life expectancy is then negative. The interpretation of the absolute value of the difference in life expectancy as the stationary-equivalent of the *MUL* is a “backward-looking” interpretation in the sense that it describes average years of life lost in a recent “death cohort” (Riffe, Schöley, and Villavicencio 2017). It provides an alternative to the forward-looking interpretation of life expectancy that refers to the hypothetical, future survival of a current birth cohort exposed to unchanging mortality conditions throughout their lifetime which becomes problematic when mortality surges are expected to be temporary. Under this alternative interpretation, the difference in life expectancy during a mortality surge does not measure changes in the survival prospects of the population that is currently living, but rather the standardized value of the average lifespan reduction of those who died during the mortality surge.

## Supporting information

Technical Appendix

## Data Availability

All data produced in the present work are contained in the manuscript.

## Acknowledgments

The author benefited from facilities and resources provided by the California Center for Population Research at UCLA (CCPR), which receives core support (P2C-HD041022) from the Eunice Kennedy Shriver National Institute of Child Health and Human Development (NICHD). I thank Hiram Beltrán-Sánchez, Noreen Goldman, Josh Goldstein, Michelle Poulin, and Tim Riffe for comments on an earlier version of this paper.

