## Supplementary material for "Interpreting Changes in Life Expectancy During Temporary Mortality Shocks": Technical Appendix

**Supplementary Information**

**Interpreting Changes in Life Expectancy**

**During Temporary Mortality Shocks**

**Part 1. Keyfitz’ approximation and the actual difference in period life expectancy at birth**

To illustrate the impact on life expectancy at birth of a hypothetical medical improvement (assuming no other change in future mortality), Keyfitz (1977) assumed the same relative improvement in age-specific death rates at all ages:

$$\frac{\left( \mu^{A}\left( a \right)-\mu^{B}\left( a \right) \right)}{\mu^{B}\left( a \right)}=\delta$$

**(S1)**

where superscripts B and A refer to the original (before improvement) and the cause-deleted (after improvement) life table respectively, and *δ* is a small negative quantity. After a Taylor expansion, Keyfitz showed that if *δ* is small enough for *δ*^2^ to be considered negligible relative to *δ*:

$$\frac{\left( e_{0}^{o} \right)^{A}-\left( e_{0}^{o} \right)^{B}}{\left( e_{0}^{o} \right)^{B}}\simeq\delta.\frac{\int_{0}^{\infty} l^{B}\left( a \right).ln\left( l^{B}\left( a \right) \right)da}{\int_{0}^{\infty} l^{B}\left( a \right)da}= -\delta.H^{B}$$

**(S2)**

where *H*^B^ is the entropy of the original life table. Goldman and Lord (1986) further show that *H*^B^ can also be written as:

$$H^{B}=\frac{\int_{0}^{\infty} d^{B}\left( a \right).\left( e_{a}^{o} \right)^{B}da}{\int_{0}^{\infty} l^{B}\left( a \right)da}$$

**(S3)**

It follows that a difference in life expectancy can be approximated as:

$$\left( e_{0}^{o} \right)^{A}-\left( e_{0}^{o} \right)^{B}\simeq-\delta.H^{B}.\left( e_{0}^{o} \right)^{B}=-\delta.\frac{\int_{0}^{\infty} d^{B}\left( a \right).\left( e_{a}^{o} \right)^{B}da}{\int_{0}^{\infty} l^{B}\left( a \right)da}.\left( e_{0}^{o} \right)^{B}= -\delta.\frac{\int_{0}^{\infty} d^{B}\left( a \right).\left( e_{a}^{o} \right)^{B}da}{\int_{0}^{\infty} d^{B}\left( a \right)da}$$

**(S4)**

The right-hand is the “e-dagger,” a concept described in Vaupel and Canudas Romo (2003):

$$\left( e_{0}^{o} \right)^{A}-\left( e_{0}^{o} \right)^{B}\simeq-\delta.\left( e_{0}^{o} \right)^{\dagger B}$$

**(S5)**

More generally, Pollard (1988: 266) showed that a difference in life expectancy can be written exactly as:

$$\left( e_{0}^{o} \right)^{B}-\left( e_{0}^{o} \right)^{A}= \int_{0}^{\infty} \left( \mu^{A}\left( a \right)-\mu^{B}\left( a \right) \right).{{}_{a}p}_{0}^{B}.\left( e_{a}^{o} \right)^{A}da$$

**(S6)**

where *μ*(*a*) denotes the mortality rate at exact age *a*, *_a_p_0_* the probability to survive from birth to age *a*, and *e_a_*^o^ life expectancy at age *a*, and the superscripts *A* and *B* refer to two different life tables (corresponding to different populations or sub-populations, or to the same population at two different times). Intuition for equation (9) can be derived from rewriting the sum as:

$$\left( e_{0}^{o} \right)^{B}-\left( e_{0}^{o} \right)^{A}=-\frac{\int_{0}^{\infty} \left( \mu^{B}\left( a \right)-\mu^{A}\left( a \right) \right).l^{B}(a).\left( e_{a}^{o} \right)^{A}da}{\int_{0}^{\infty} \mu^{B}\left( a \right).l^{B}(a)da}$$

**(S7)**

To tie Pollard’s more general approach to Keyfitz’ approximation, in the case of identical relative changes in mortality rates at all ages expressed in equation (S1), equation (S7) becomes:

$$\left( e_{0}^{o} \right)^{B}-\left( e_{0}^{o} \right)^{A}=\delta.\frac{\int_{0}^{\infty} \mu^{B}\left( a \right).l^{B}\left( a \right).\left( e_{a}^{o} \right)^{A}da}{\int_{0}^{\infty} \mu^{B}\left( a \right).l^{B}\left( a \right)da}=\delta.\frac{\int_{0}^{\infty} d^{B}\left( a \right).\left( e_{a}^{o} \right)^{A}da}{\int_{0}^{\infty} d^{B}\left( a \right)da}$$

**(S8)**

As shown in the last ratio, the difference in life expectancy is then proportional to a weighted average of life expectancies after the mortality change, with the weights provided by life table decrements in the original table (before the change). The exact equation (S8) is very similar to the approximation in equation (S4). If relative changes in mortality are the same at all ages and relatively small, the approximation in equation (S4) shows that original life expectancies (that include the “deleted” cause of death) can be used instead of the life expectancies after the deletion. (Equation (S4) could also be derived directly from equation (S8) through a Taylor expansion).

**Part 2. The MUL and other indicators of premature mortality**

The *MUL* is an average number of years of life lost to excess mortality, *YLL^E^*, per death in population *B*. Averaging instead *YLL^E^* over the number of excess deaths, *D^E^*, would indicate the average number of years of life lost per excess death in population *B*. To express this, the *MUL* can be written as:

$$MUL=\frac{D^{E}}{D^{B}}. \frac{\int_{0}^{\infty} D^{E}(a).\left( e_{a}^{o} \right)^{A}da}{D^{E}}$$

**(S9)**

The first ratio expresses the relative incidence of excess and all-cause mortality, frequently measured via the *P-score*:

$$P= \frac{\int_{0}^{\infty} \left( \mu^{B}\left( a \right)-\mu^{A}\left( a \right) \right).N^{B}(a)da}{\int_{0}^{\infty} \mu^{A}\left( a \right).N^{B}(a)da}$$

**(S10)**

In the case of identical relative increases in mortality at all ages, the *P-score* equals the scalar *δ* in equation (S1), which is then positive. The first ratio in equation (A9) can be written as a function of the *P-score*:

$$\frac{D^{E}}{D^{B}}= \frac{\int_{0}^{\infty} \left( \mu^{B}\left( a \right)-\mu^{A}\left( a \right) \right).N^{B}(a)da}{\int_{0}^{\infty} \mu^{B}\left( a \right).N^{B}(a)da}= \frac{\int_{0}^{\infty} \left( \mu^{B}\left( a \right)-\mu^{A}\left( a \right) \right).N^{B}(a)da}{\int_{0}^{\infty} \mu^{A}\left( a \right).N^{B}(a)da}.\frac{\int_{0}^{\infty} \mu^{A}\left( a \right).N^{B}(a)da}{\int_{0}^{\infty} \mu^{B}\left( a \right).N^{B}(a)da}=\frac{P}{1+P}$$

The *MUL* can then be expressed as:

$$MUL= \frac{P}{1+P}.\frac{\int_{0}^{\infty} D^{E}(a).\left( e_{a}^{o} \right)^{A}da}{D^{E}}$$

**(S11)**

In the literature on premature mortality, *YLL^E^* is often related to the total size of the population instead. To express this, the *MUL* can also be written as:

$$MUL=\frac{N^{B}}{D^{B}}. \frac{\int_{0}^{\infty} D^{E}(a).\left( e_{a}^{o} \right)^{A}da}{N^{B}} = \frac{1}{{CDR}^{B}}. \frac{\int_{0}^{\infty} D^{E}(a).\left( e_{a}^{o} \right)^{A}da}{N^{B}}$$

**(S12)**

where *CDR^B^* is the Crude Death Rate and *N^B^* is the population size in population *B*.

Both terms of the product in equation (S12) can be standardized using an “external” standard population distribution, *N^S^*(*a*). After standardization, the first ratio would become the inverse of the classic age-standardized crude death rate, *ASCDR*, in population *B* (Preston, Heuveline, and Guillot 2001):

$${ASCDR}^{B}= \frac{\int_{0}^{\infty} N^{S}(a).\mu^{B}\left( a \right)da}{\int_{0}^{\infty} N^{S}(a)da}$$

**(S13)**

The second ratio can be written as:

$$\frac{\int_{0}^{\infty} D^{E}(a).\left( e_{a}^{o} \right)^{A}da}{N^{B}}=\frac{\int_{0}^{\infty} N^{B}\left( a \right).\left( \mu^{B}\left( a \right)-\mu^{A}\left( a \right) \right).\left( e_{a}^{o} \right)^{A}da}{\int_{0}^{\infty} N^{B}(a)da}$$

**(S14)**

It appears as a weighted average of the *YLL^E^* rates at age *a* (Martinez et al. 2019), *YLLR^E^*(*a*):

$${YLLR}^{E}\left( a \right)= \frac{{YLL}^{E}\left( a \right)}{N^{B}(a)}=\frac{D^{E}\left( a \right).\left( e_{a}^{o} \right)^{A}}{N^{B}(a)}=\frac{\left( \mu^{B}\left( a \right)-\mu^{A}\left( a \right) \right).N^{B}(a).\left( e_{a}^{o} \right)^{A}}{N^{B}(a)}=\left( \mu^{B}\left( a \right)-\mu^{A}\left( a \right) \right).\left( e_{a}^{o} \right)^{A}$$

**(S15)**

In equation (S14), the weights are provided by the population distribution. If we substitute instead the standard population distribution, we get the age standardized *YLL^E^* rate, *ASYR^E^*:

$${ASYR}^{E}= \frac{\int_{0}^{\infty} N^{S}(a).{YLLR}^{E}\left( a \right)da}{\int_{0}^{\infty} N^{S}(a)da}$$

**(S16)**

The ratio *ASYR^E^*/*ASCDR^B^* is thus an externally standardized *MUL*, whereas the absolute value of the difference in life expectancy is an internally standardized *MUL*.
